# Epidemiological pattern of trauma among children 0 - 9 years in Cameroon

**DOI:** 10.1101/2024.09.19.24314028

**Authors:** Odette Kibu, Sithombo Maqungo, Georges Nguefack-Tsague, Sithombo Maqungo, Dickson Shey Nsagha, Fanny N. Dissak Delon, Darwin Touko, Elvis Asangbeng Tanue, Rasheedat Oke, Sandra I. McCoy, Sabrinah Ariane Christie, Catherine Juillard, Alain Chichom Mefire

**Affiliations:** Department of Public Health and Hygiene, University of Buea, Faculty of Health Sciences, Buea, Cameroon; Division of Global Surgery and Division of Orthopeadic Surgery, University of Cape Town and Groote Schuur Hospital, South Africa; Department of Public Health, Faculty of Medicine and Biomedical Sciences, University of Yaoundé I, Cameroon; Data Science Center for the Study of Surgery, Injury, and Equity in Africa (D-SINE-Africa), University of Buea, Buea, Cameroon; Department of Surgery, Program for the Advancement of Surgical Equity, University of California Los Angeles, Los Angeles, California, USA; Division of Epidemiology, University of California Berkeley, Berkeley, USA; Department of Surgery, University of Buea Faculty of Health Sciences, Buea, Cameroon

**Keywords:** Cameroon, Children, Epidemiological pattern, Injury, Trauma

## Abstract

**Background:** In low- and middle-income countries, trauma is the leading cause of death among youth and it is also a major cause of disability. Globally, more than 1,600 children and adolescents below the age of 19-years die every day from preventable injuries. Traffic-related injuries, falls, sports-related injuries, assaults, burns, and drownings are the most commonly reported causes of traumatic mortality among children. The mechanism of injury is always diverse in different contexts due to the differences in social determinants of health.

**Objectives:** To determine the epidemiological pattern of trauma among children 0 – 9 years in Cameroon.

**Methods:** This is a retrospective analysis of prospectively collected data from the Cameroon Trauma Registry currently running in 10 pilot sites across seven of the ten regions of Cameroon. We retrieved data for all children aged 0 - 9 years from June 2022 to August 2023. Data was analyzed with respect to the demographics, injury characteristics and outcomes.

**Results:** Of the 5,439 patients captured in the trauma registry, 267 (4.9%) were children aged 0-9 years. Over 50% (152/267) of the patients were males with 35% (93/267) from rural settings. The top injury mechanism was road traffic injuries (RTI) [137(52.1%)]. These injuries occurred on the streets [142(53.4%)] during leisure activities [205(78.5%)]. Majority of children [104(39%)] involved in RTI injuries were pedestrians and no prehospital care offered to 216(82.8%) of injured children. A total of 39 (16.9%) were discharged with major disability, 111 (48.1%) had limited ability to move and 5 (2.2%) demised or succumbed to their injuries. There was a significant association between injury activity and gender (P = 0.006). Unlike the females, majority of the males were discharged with major disability [21 (53.8%)].

**Conclusion:** This preliminary analysis highlights the burden of trauma among children aged 0 – 9 years and its contribution to the proportion of disabled persons in Cameroon. Leisure activities on the streets increased the number of pediatric injuries especially among males. It is therefore imperative to put in place or reinforce environmental interventions to reduce the burden of pediatric injuries.

## 1. Introduction

Childhood trauma has increasingly become a global public health concern in both developed and developing countries (1) and approximately 90% of these injuries are unintentional and occur in mostly low- and middle-income countries (2). According to the 2008 WHO/UNICEF World report on child injury prevention, injury and violence are major killers of children throughout the world, responsible for approximately 950, 000 deaths in children and young people under the age of 18 years each year (3). That is more than 100 children dying from preventable death every hour of every day (3). Unintentional injuries, including road traffic injuries, falls, burns, poisoning and drowning, account for almost 90% of these cases (3). The burden associated with childhood trauma is worse in Sub-Saharan Africa where children are approximately 5 times more likely to die from trauma compared to children in developed countries (4). Trauma in children is not only a major public health issue, but it has significant effects on the social and economic developments of a country.

In sub Saharan Africa (SSA), injuries have now become the third leading cause of death (5) (6) and the region has the world’s highest rate of unintentional injury deaths among children in particular aged 1-4 years at 100.5 per 100,000 (7). Research has shown males have a higher risk of injury compared to females (8). In addition to the deaths caused by injury, many more children are left with some form of disability or disfigurement that impedes their full integration in social and economic life (6). Over a third of these injury deaths can be prevented by access to adequate trauma care (9).

Epidemiological studies on childhood trauma in Cameroon are generally lacking and as a result, few injury prevention and control programs that are based on evidence exist in the country. Previous studies have focused on specific forms of trauma such as road traffic injuries, burns or have been sub-national in scope (10) (11) (12) (13). Comprehensive injury surveillance programs for creation of a national injury prevention policy are still lacking. Moreover, it is not known what the current burden of trauma on children of different sex and age groups in Cameroon is. This study aims to investigate the epidemiological pattern of trauma among children aged 0-9 years in Cameroon. By identifying and documenting the current local patterns of childhood trauma in Cameroon, it is expected that the findings of this study together with other ongoing efforts will provide a strong scientific foundation and momentum for the support of an injury prevention action plan/program in the country. This will ultimately help the Ministry of Public Health and other partner organizations to utilize the best available data in their efforts to protect and safeguard the well-being of children in Cameroon.

## 2. Methods

### 2.1. Study design

This was retrospective analysis of prospectively collected data from the Cameroon Trauma Registry (CTR) from June 2022 to May 2023 (12 months). The CTR is implemented in 10 hospital sites across seven of the ten regions of Cameroon. These hospitals are: Laquintinie Hospital of Douala, (HLD), Limbe Regional Hospital, (HRL), Pouma Catholic Hospital, (HCP), Edea Regional Hospital, (HRE), Emergency and Reanimation Center of Yaounde, (CUY), Bafoussam Regional Hospital (HRB), Maroua Regional Hospital (HRM), Bafia district hospital (HDB), Kribi District hospital (HDK), and Bertoua Regional Hospital (BRH).

This registry collects prospective information about demographics, nature and context of the injury, delay in accessing care, clinical course and outcome of care.

### 2.2. Study population

All trauma patients with the age 0 – 9 years presenting to the Emergency Department of the 10 hospitals with an acute traumatic injury as defined by the WHO, were extracted from the CTR and included in our study. CTR includes patients with acute traumatic injury who presented for care within 2 weeks of the injury date, trauma patients who were formally admitted to the hospital as in-patients, or who were transferred to other health facilities after being kept under observation in the emergency department for over 24 hours.

### 2.3. Data collection

The implementation of the CTR started on 1^st^ June 2022 and it currently being implemented still the year 2026. Trained data collectors for the CTR were placed at the emergency department of the 10 hospitals to collect data prospectively from trauma patients. Eligible trauma patients were recruited into CTR and followed-up in the hospital until the date of disposition (discharged, died, transferred or left against medical advice). CTR uses a structured questionnaire to collect data daily which is later entered into Research Electronic Data Capture, (REDCap) which is a Health Insurance Portability and Accountability (HIPAA)-compliant, secure web application for building and managing online surveys and databases. A well-structured data extraction guide was used to extract data from the trauma registry about pediatric trauma from the 1^st^ of June 2022 to 31^st^ August 2023. The data was related to demographics, injury characteristics, injury mechanism and outcomes. Injuries were categories into intentional and unintentional (these are subset of injuries for which there is no evidence of predetermined intent).

### 2.4. Ethical considerations

Participation in the CTR was voluntary and verbal consent was obtained from the guardians of the children. Each participant’s form had a section that documented that verbal consent was obtained before data collection. Access to patients’ data by the authors was de-identified from the authors during and after data collection CTR does not collect data from minors without the consent of the guardian. Ethical approval for the CTR was obtained from the University of California, Los

Angeles Institutional Review Board (#19-000086, approved on 13^th^ June 2019), from the Division of Health Operation Research at the Ministry of Public Health (D30-923/L/MINSANTE/SG/DROS, approved on 8^th^ August 2022), from the Faculty of Health Sciences Institutional Review Board of the University of Buea (2021/1506-07/UB/SG/IRB/FHS, approved on 30^th^ September 2021) and from National Ethics committee (N° 2018/09/1444/CE/CNERSH/SP, approved on 25^th^ March 2022).

### 2.5. Data Analysis

The CTR data entered into REDCap was exported to a Comma Separated Value (CSV) file for analysis in the R software 4.2.1. All entered data were verified for completeness and accuracy and later on cleaned for analysis. Univariate analysis for categorical data were presented as frequency and percent distribution and bivariate analysis was done using the fisher exact test to measure association of dependent variables and sex

## 3. Results

### 3.1. Socio demographic characteristics of the study population

Out of the 5,439 trauma patients included in the Cameroon trauma registry (CTR), a total of 267 (4.9%) participants were aged 0 – 9 years with majority beings males [152 (56.9%)] with a mean age of 4.9 ± 2.7 years. Majority of the participants were from the Bertoua regional hospital [68 (25.5%)] and resident in urban areas [173 (65%)] as shown in Table 1. Primary level of education was the category with highest number of participants [152(56.9%)].

**Table 1:**
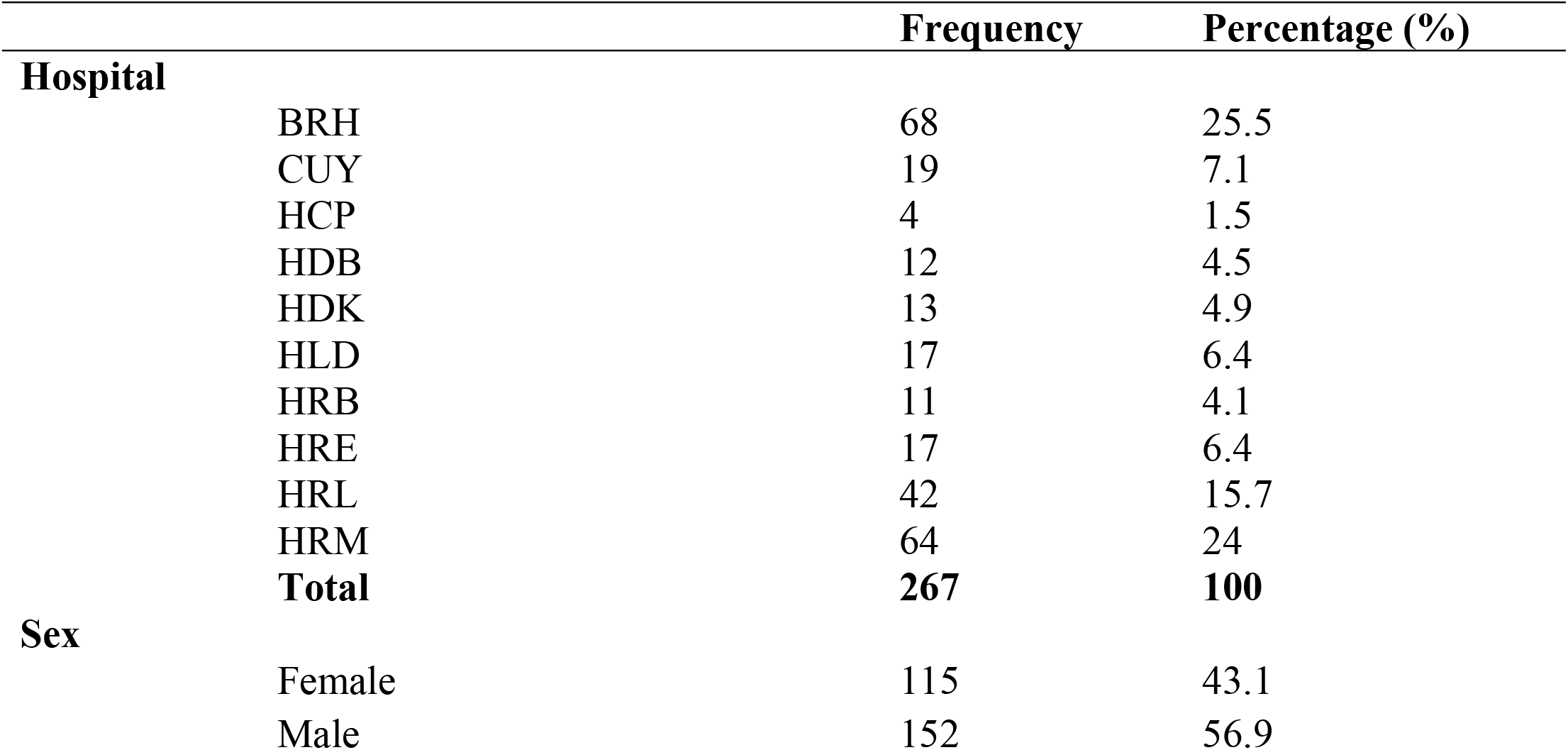
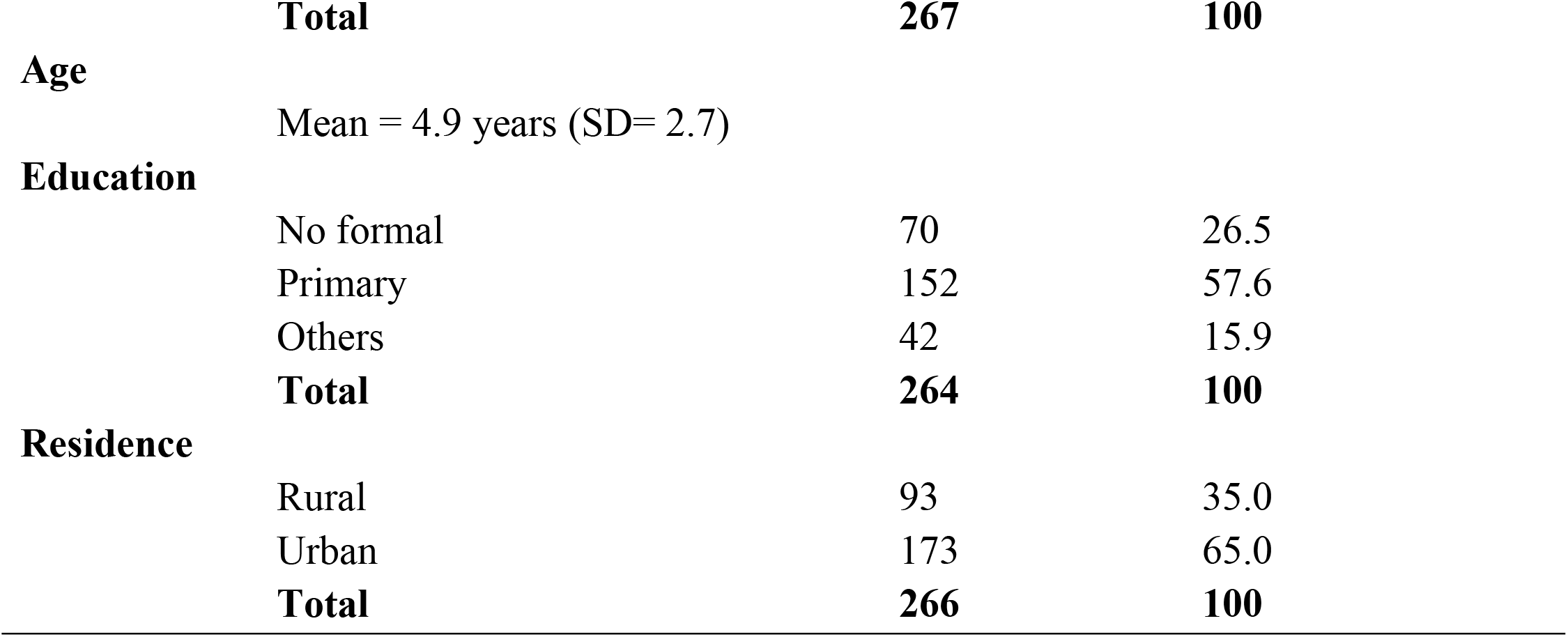
Socio-demographic characteristics of study population.

|  | Frequency | Percentage (%) |
| --- | --- | --- |
| **Hospital** |  |  |
| BRH | 68 | 25.5 |
| CUY | 19 | 7.1 |
| HCP | 4 | 1.5 |
| HDB | 12 | 4.5 |
| HDK | 13 | 4.9 |
| HLD | 17 | 6.4 |
| HRB | 11 | 4.1 |
| HRE | 17 | 6.4 |
| HRL | 42 | 15.7 |
| HRM | 64 | 24 |
| **Total** | **267** | **100** |
| **Sex** |  |  |
| Female | 115 | 43.1 |
| Male | 152 | 56.9 |

|  |  |  |  |
| --- | --- | --- | --- |
|  | **Total** | **267** | **100** |
| **Age** | Mean = 4.9 years (SD= 2.7) |  |  |
| **Education** | No formal | 70 | 26.5 |
|  | Primary | 152 | 57.6 |
|  | Others | 42 | 15.9 |
|  | **Total** | **264** | **100** |
| **Residence** | Rural | 93 | 35.0 |
|  | Urban | 173 | 65.0 |
|  | **Total** | **266** | **100** |

### 3.2. Injury characteristics among the study population

Overall, majority of the children had injury during leisure activities [205(78.5%)] of which [142 (53.4%)] occurred on the streets. Road traffic injury was the main cause of injury [137 (52.1%)] followed by falls [52 (19.8%)] as shown in Table 2.

**Table 2:** Injury characteristics of the study population.

|  | **Frequency** | **Percentage (%)** |
| --- | --- | --- |
| **Injury activity** |  |  |
| Leisure/play | 205 | 78.5 |
| Traveling | 23 | 8.8 |
| Domestic activity | 19 | 7.3 |
| Education | 6 | 2.3 |
| Work | 2 | 0.8 |
| Other | 6 | 2.3 |
| **Injury location** |  |  |
| Road | 142 | 53.4 |
| Private house/home | 104 | 39.1 |
| School area | 6 | 2.3 |
| Farm | 6 | 2.3 |
| Water body | 3 | 1.1 |
| Others | 5 | 1.9 |
| **Role of patient** |  |  |
| Pedestrian | 104 | 75.9 |
| Mototaxi | 14 | 10.2 |
| Motorcycle | 8 | 5.8 |
| Bus | 5 | 3.6 |
| Private Car | 4 | 2.9 |
| Others | 2 | 1.6 |
| **Counterpart** |  |  |
| Mototaxi | 75 | 55.1 |
| Vehicle | 26 | 19.1 |
| Motorcycle | 11 | 8.1 |
| Truck/Camion | 9 | 6.6 |
| None | 13 | 9.6 |
| Other | 2 | 1.5 |

An analysis of the 137 (52.1%) children involved in RTI as shown in Figure 1 showed that, among this number a majority [98 (71.5%)] were involved in leisure activities and most of the injury victims were pedestrians [104 (75.9%)]. Majority of the children were hit by a motor taxi [75(55.1%)]. Among 10 children who were involved in a car or bus injury only 1 (0.1%) used a car seat.

**Figure 1:**
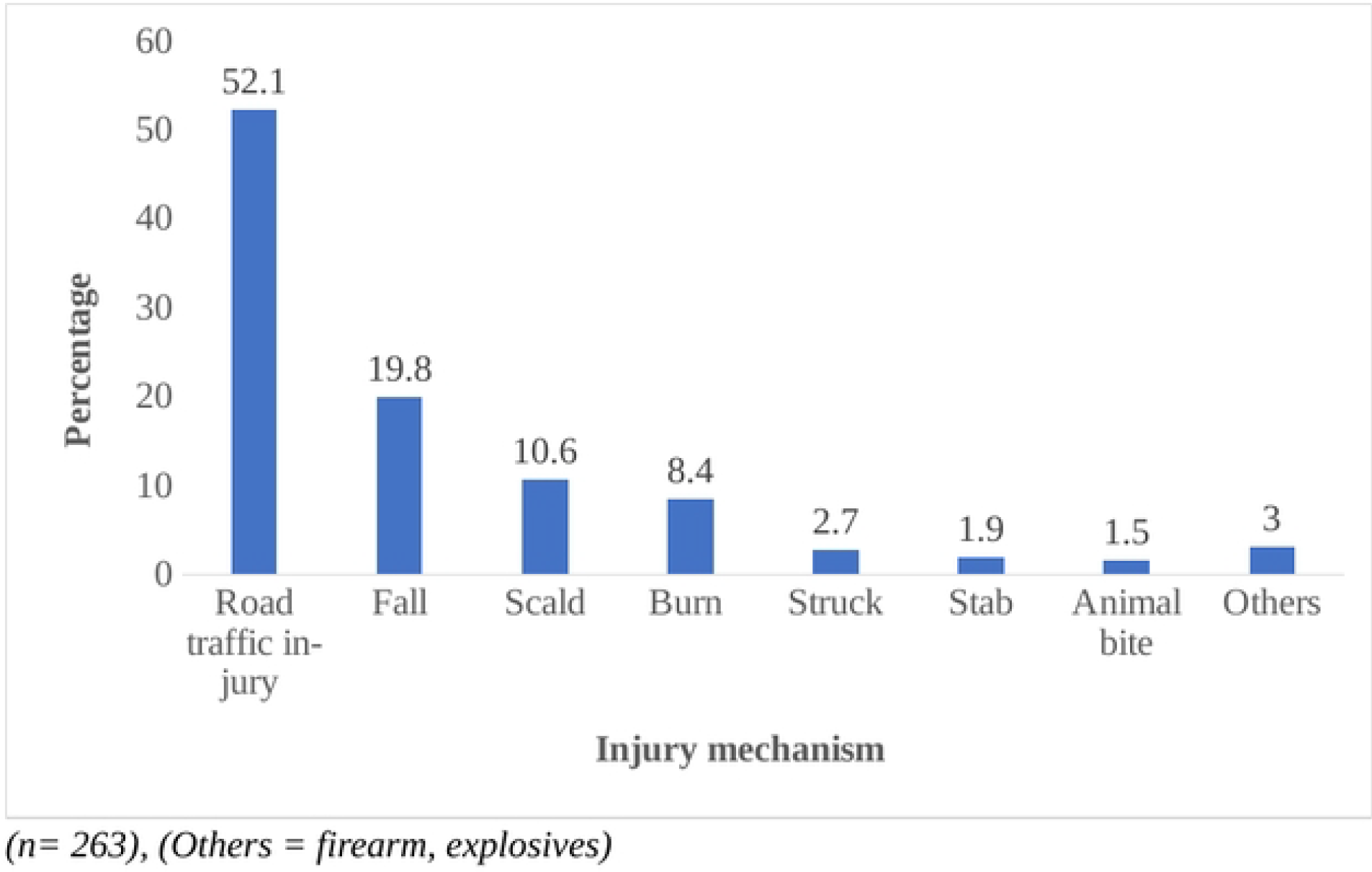
Proportion of children with different injury mechanism in the 10 hospitals site over 12 months period

Overall, 98.5% were involved in unintentional injury, no scene care offered to 216 (82.8%) of the children involved in any form of injury as shown in Figure 2.

**Figure 2:**
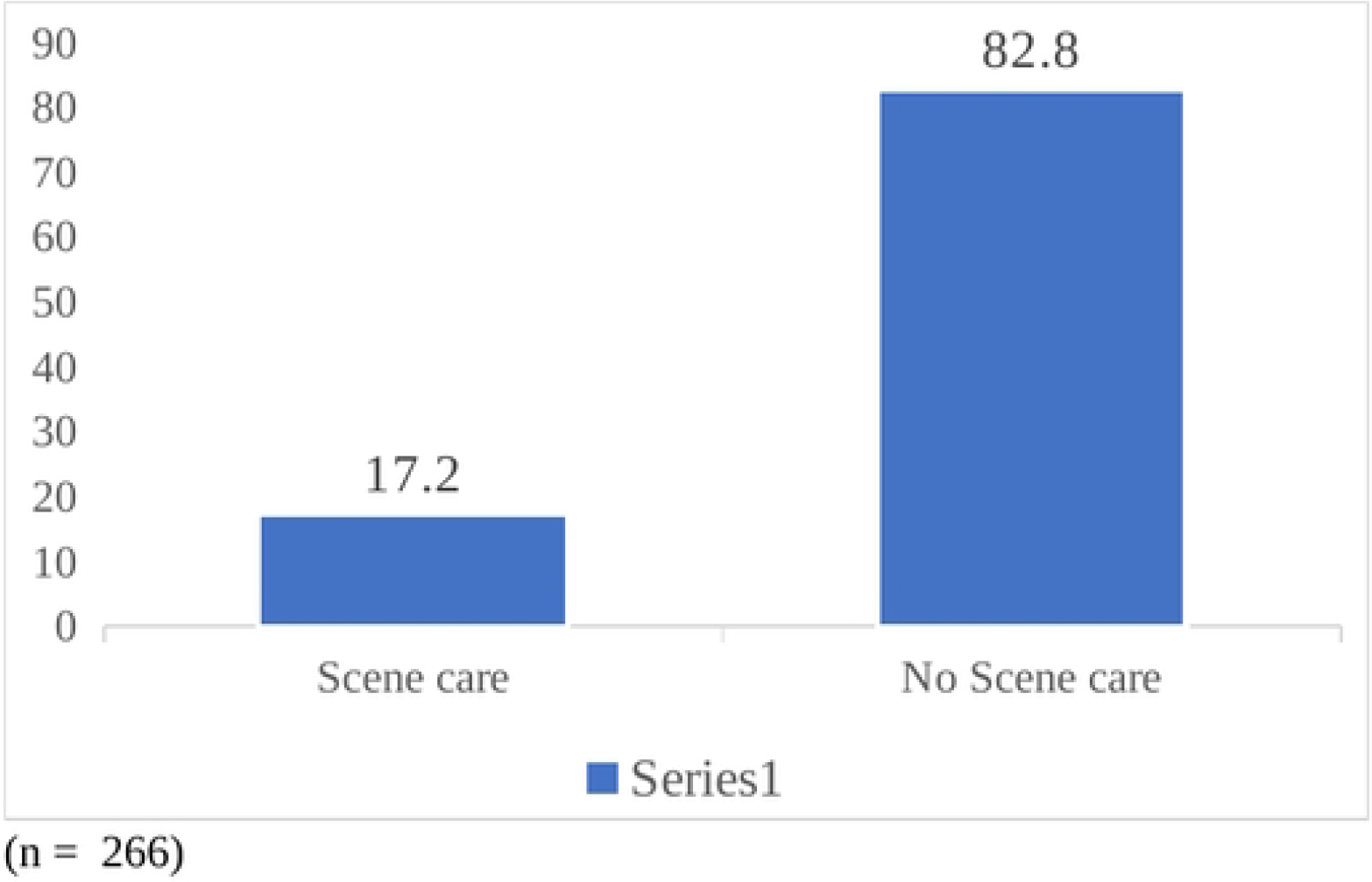
Proportion of children who received scene care from the 10 hospitals site over 12 months period

Figure 3 shows the outcome of the patients after hospitalization whereby 2.2% died, 16.9% left the hospital with a major disability while 48.1% had minor disability with a limited ability to move.

**Figure 3:**
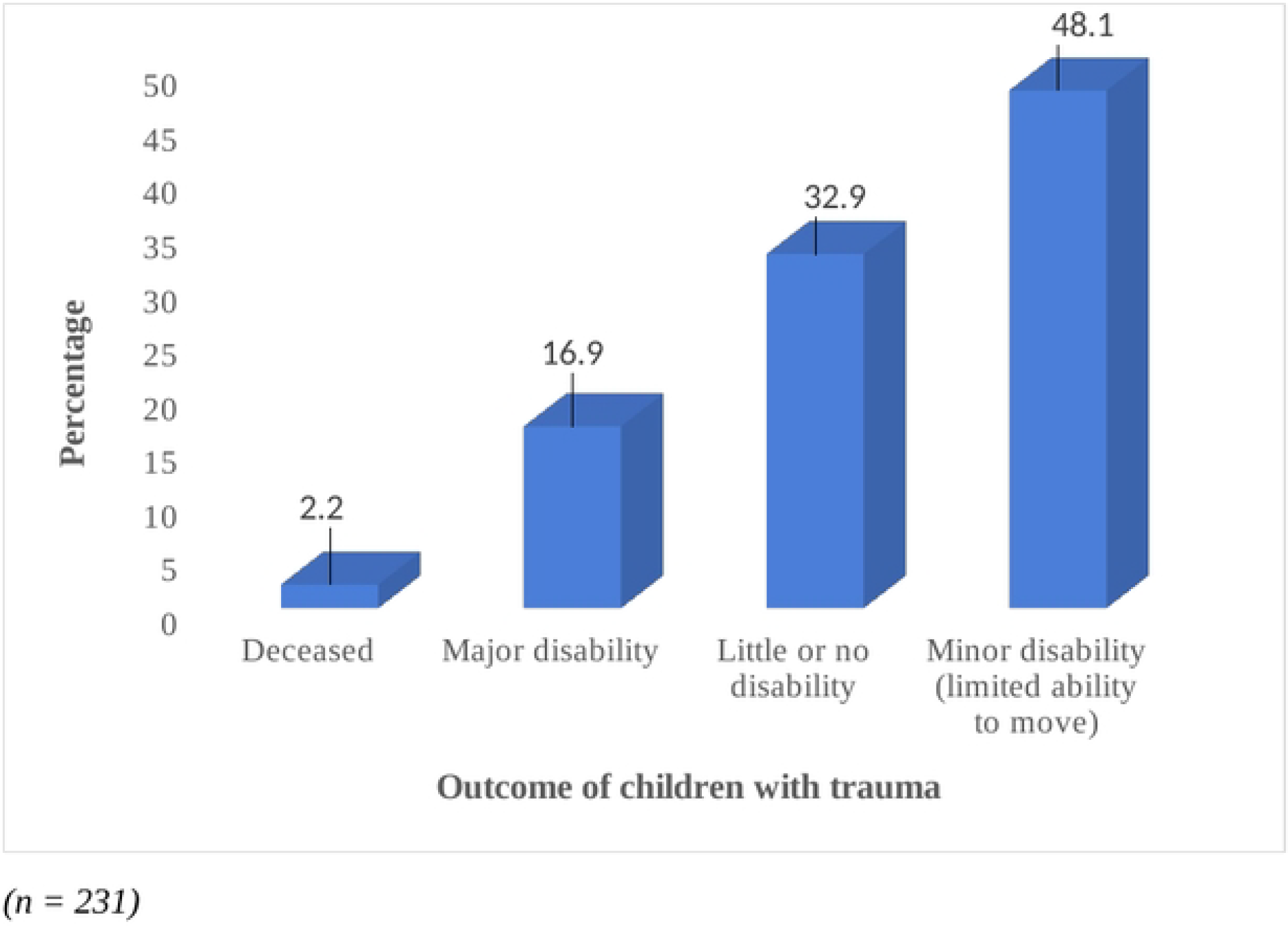
Outcome of the patients after hospitalization 10 hospitals site over 12 months period

### 3.3. Association between injury characteristics and gender

Analysis of the results in Table 3 showed that majority of the injured participants were males who were mostly involved in leisure activities by the road side. Compared to the males, females were mostly injured during domestic activities [12 (63.2%)]. Majority of males had Injury Severity Score (ISS) of 3 [11 (64.7%)]. Further analysis showed that there was an association between gender and injury activity (P = 0.006)

**Table 3:**
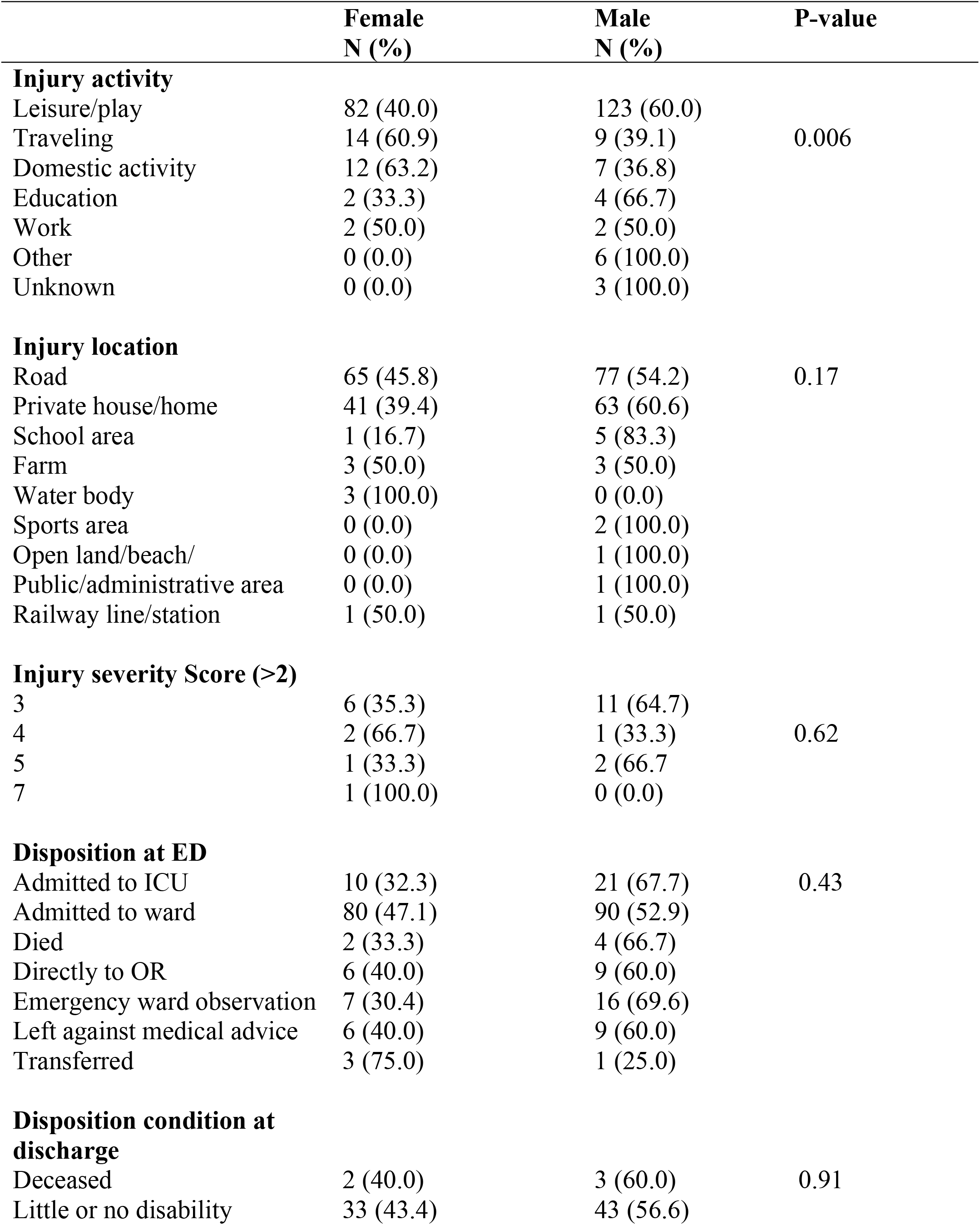
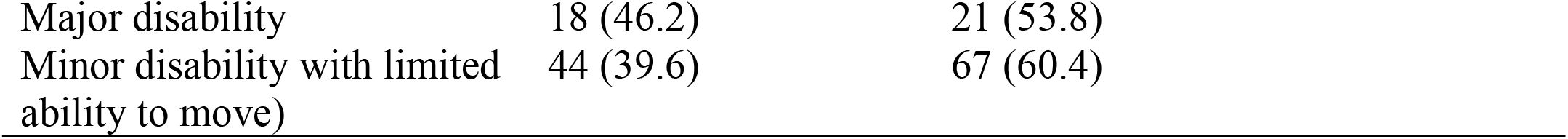
Association between injury characteristics and gender.

At the Emergency Unit, the study showed that mostly males were either admitted to Intensive Care Unit (ICU) [21 (67.7%)] or sent directly to the Operating Room (OR) [9 (60%)] or emergency ward observation [16 (69.6%)] or to the ward for further observation and treatment [90 (52.9%)]. Unlike the females, majority of the males were discharged with major disability [21 (53.8)]. However, there was no association between sex and the disposition of the children either at the emergency or ward as show in Table 3.

## 4. Discussion

In this study we observed that RTI are the main cause of injury in children and these injuries are unintentional leading to a majority of disabled children.

A proportion of 98.5% were involved in unintentional injury and most of which occurred on the streets. Research has shown that unintentional injury accounts for one-third of deaths in children and adolescents each year, primarily from motor vehicle crashes (14). In a study conducted in Rwanda, road traffic injuries constituted 15% of all child injuries and was the most common unintentional injury in children over five years (15). Consistent with other studies, we found that most injured children were pedestrians (16) (17). Gong et al found out that preschool children under the age of 6 years old account for approximately 64.6% of unintentional injuries in the emergency department (18).

This might be due to the children playing on the streets without supervision and no environment intervention. It is important to take into consideration the behaviors of children when developing prevention measures especially on the roads.

Injuries are also a primary cause of disability, which can have a long-lasting effect on the lives of the children, namely in relationships, learning, and games. Road traffic injury was the major cause of injury affecting 52.1% of the children in this study. Other studies in Asia have shown that road traffic injuries are one of the five leading causes of disability for children (19). According to these surveys, the rate of permanent disability among children aged 1 to 17 years injured as a result of a road traffic crash was 20 per 100 000 children. As a result of the injuries children are likely to miss schools or are hospitalized. Contrary to our study, other authors have shown that unintentional falls are the leading cause of non-fatal injuries in Emergency Department visits among children under the age of 14 years old (20) (21) (22).

Results from this study showed that majority of the children left the hospital with a disability with 16.9% having a major disability. The disability in children could have short-term and long-term impact on these young patients’ functionality and quality of life. Most of the injuries experienced by children are preventable and yet they are so common due to various factors such as: lack of legislation or enforcement to promote child safety, lack of environmental modifications to create safe play areas and housing, low prioritization of child safety, and inadequate adult supervision (23). Interventions such as environmental modification and improved surveillance systems tailored towards reducing child injuries will go a long way to reduce disabilities and preserve the joy of childhood (24).

Males were mostly injured compared to females. They made up a higher proportion of those who left the wards with disability. In line with other studies, Adam and Matheny showed that boys had a higher number of injuries than the girl co-twins. This was due to higher activity level, less focused attention, more noise-confusion in the household, less family organization-control, and mothers’ being less sociable or more preoccupied. Their results also suggest that higher injury rates for boys accrue from a range of psychosocial and environmental influences wider than that identified for girls (8). In Ghana, a study conducted by Abantanga and Mock showed that boys sustained higher injury rates in all age groups than girls, with an overall rate of, 136/100000 children as compared to 92/100000 for girls (25).

### Strengths and Limitation

This study was able to capture data from 7 out of the 10 regions of Cameroon over 1 year period. The study sites chosen receive a representative number of patients from its region. Our study excluded unconscious patients without a guardian to consent. Thus, a selection bias might have been introduced as unconscious patients embody a specific subgroup with different characteristics.

## Conclusion

This study showed the most common causes of unintentional injury among children aged 0-9 years. These injuries occur when they are involved in leisure activities by the road. Major disability was found in 16.9% of the population and the males were the most affected. This study strongly recommends safety behaviours, environmental interventions and parent/guidance supervision as possible interventions to reduce the injuries in this group of the population who are most at risk of sustaining trauma.

## Data Availability

Data cannot be shared publicly because because it is hosted by the University of Buea. Data are available from the University of Buea Institutional Data Access for researchers who meet the criteria for access to confidential data.The data underlying the results presented in the study are available upon request to the corresponding author

## Acknowledgments

We acknowledge the National Institute of Health and all participants that took part in the Cameroon Trauma registry.

## Declaration of Interest Statement

The authors declare that they have no known competing financial interests or personal relationships that could have appeared to influence this research.

## Notes

### Competing Interest Statement

The authors have declared no competing interest.

### Funding Statement

Yes

### Author Declarations

Ethical approval for the CTR was obtained from the University of California Los Angeles Institutional Review Board (#19-000086), from the Cameroonian National Ethics Committee (N° 2018/09/1444/CE/CNERSH/SP), from the Faculty of Health Sciences Institutional Review Board of the University of Buea (2021/1506-07/UB/SG/IRB/FHS).

